# Women Empowerment and its Effect on Female Genital Mutilation in Sub-Sahara Africa: A study using Recent Demographic Health Surveys

**DOI:** 10.1101/2024.09.03.24313034

**Authors:** Munawar Harun Koray, John Mugisha

## Abstract

**Background:** Women empowerment can enhance and improve women decsions and will power to negotiate and stand against sensitive issues that affect women’s health and sexuality. Practices such as Female Genital Mutilation (FGM) are generally sensitive issues that requires education and holistic interventions to enable its eradication, particularly in sub-Sahara Africa (SSA), where the practice is still pervasive. This study investigated the effect of women empowerment, using the Survey-base Women Empowerment (SWPER) Global index, on FGM in selected countries in SSA.

**Methods:** The study employed cross-sectional design using the most recent Demographic Health Survey (DHS) data from 10 SSA countries. A total of 49501 women who were either married or living with a partner and have a daughter age 0 – 14 years who is either circumcised or not. Pearson’s chi-square test was used to examine the association between women empowerment and daughter with FGM, and countries. Bivariate and multivariate logistic regressions were used to examine the predictors of FGM at a significance level of p ≤ 0.05 and 95% confidence interval. Complex survey design was considered in the analysis.

**Results:** 49,501 participants were included in the study and 47.96% exhibited a positive attitude towards violence. Empowerment in SWPER domains were significantly associated with FGM (p < 0.0001), with Kenya and Tanzania exhibiting high levels of women empowerments and low prevalence of FGM. Medium and highly-empowered mothers had significantly lower odds of having a child with FGM in the bivariate regression models (p < 0.0001), compared to mothers with low empowerment levels. When adjusted for confounders, increasing age is associated with higher odds of FGM with mothers age 45-49 mothers having more than 4 times higher odds (AOR: 4.265, 95%CI: 3.466 – 5.248). Muslim mothers also had higher odds of having a child with FGM (AOR: 6.046; 95%CI: 5.605 – 6.521) compared to Christian mothers. An increase in the wealth index was also a protective factor against FGM (p < 0.0001). Circumcised mothers were more likely to have their female child circumcised (AOR: 5.527; 95%CI: 5.113 – 5.975) and female household heads were found to be protective factors against FGM (AOR: 0.846; 95%CI: 0.774 – 0.925).

**Conclusion:** The study highlights a connection between women’s empowerment and FGM prevalence in SSA, highlighting the need to promote women’s autonomy and reject violence. Traditional norms and cultural support for FGM persist, particularly in Western Africa. Targeted empowerment initiatives, education, and strengthening legal frameworks can help reduce FGM prevalence.

## Introduction

Female genital mutilation or cutting (FGM) continue to be a public health concern, particularly in sub-Saharan Africa, due to its impact on the physical and mental wellbeing of women and girls who fall victim to the practice. It involves the partial or complete excision of the female external genitals or any additional harm to the female genitals (1–4). This causes significant harm, especially the long-term effects of scarring, discomfort, bleeding, infection, urinary issues, cysts, birthing complications, and higher rates of neonatal and maternal mortality (1, 4, 5). FGM is considered a violence targeting women and girls leading to gender inequity (1, 5) and it violates the rights of children and constitutes child abuse (5, 6).

FGM has four types: Clitoridectomy also known as Type 1, involving part or complete excision of the clitoris or rarely, excision of the prepuce and Excision also known as Type 2, which involves fully extracting the clitoris with either labia minora and majora or the minora only (1, 7–9). Infibulation also known as Type 3, involves narrowing the establishment of a seal over the vaginal opening of the vaginal entrance through surgical suture of either labia minora or majora and can involve clitoris removal whereas Type 4 consists of other dangerous procedures on genitals, which are not medical such as piercing, sharp penetrating, incision, scraping, and cauterizing (1, 7–9).

FGM is an illegal activity in many parts of the world: The United Nations (UN) and World Health Organization prohibit it (4); the Association of American Medical Doctors (10) and the Global Federation of Obstetricians and Gynecologists (11) all stand against any form of FGM. The international community has for the past two decades, strived to eliminate FGM (12), the International Conference for Population Development, which convened governments in 1994, called them to eradicate FGM (13). Every 6 February, the UN recognizes the International Day of Zero Tolerance for FGM, with resolutions from 2012 and 2014 aiming to intensify global efforts to curb FGM (5).

A section of the Sustainable Development Goals (SDG) aim at eradicating FGM by 2030 (13, 14), and twenty-four African nations of thirty nations that practice FGM, have officially banned FGM (4, 5, 12). Albeit the action targets, without immediate and rapid responses to this public health issue, approximately, 35% of newborns worldwide will be born in countries with environments that still practice FGM (4, 5, 12) putting them at risk of FGM and other related injuries and diseases.

Worldwide, at least 230 million females have undergone FGM, with Africa being the most prevalent, with around 144 million (15). The United Nations International Children’s Emergency Fund (UNICEF) highlights that a third of children under nineteen years are the most exposed to mutilations (5) and 3.6 million females are prone to undergoing FGM annually (16, 17). The tools used in excisions are; unsterile knives, scissors, glasses, razor blades, hot materials, and clippers, among others (18, 19). Most of the time, these mutilations are done in an unhygienic manner with repeated use of the same tools (20), which poses a great risk to infectious diseases.

Women’s intent to FGM has a vast impact on the community and empowered women can raise their voices and advocate for young and aged women to abstain from FGM practices (21). This happens as women have been misrepresented, leading to barriers weakening them and leaving a significant gap in gender disparity (22). Indicators like religion, wealth index, education level, age, marital status, health status, and justifying sexual assaults are the components used in reimagining the idea of empowering women (23–26). Several Findings have demonstrated that empowering women can enhance decisions and improve their negotiation on sensitive areas that affect the population (27–30).

Over the recent years, women’s empowerment has slightly increased with education still lagging the most affected countries are in the western part of Africa, and Southern African countries mostly improving (31). Sub-Saharan African Countries and their women’s role in fighting or scrambling to end FGM practices have raised concerns as to which level of empowering a woman might be linked to exercising or eliminating FGM practices (23). Although there are several measures of women empowerment, the Survey-based Women Empowerment (SWPER) model, not without criticism, has proven to be the most robust and context based in assessing women empowerment using survey data in the SSA region (32–38).

Earlier studies conducted in SSA regarding FGM did not either fully explore women empowerment and its impact on FGM or failed to scrutinize the socio-cultural and geographical context within SSA that influences both women empowerment and FGM (39–43). Also, although the study by Coll, et. al., (2021) used the SWPER model to assess women empowerment and FGM in SSA, it did not assess a combined empowerment level for all domains of SWPER (41).

This current study seeks to contribute to policy and literature by investigating the impact of women empowerment on FGM in SSA using SWPER model and the most recent Demographic Health Survey (DHS) data. The study will also take keen considerations of the geographical, religious and socio-cultural factors that could predict the practice of FGM in the diverse regions of SSA.

## Methods

### Study Design and Data Source

This research utilized a cross-sectional design, drawing on data from the latest Demographic and Health Surveys (DHS) conducted in 10 Sub-Saharan African countries between 2010 and 2023. The DHS, a nationally representative survey, is carried out every five years in over 85 low- and middle-income countries globally (44). Participants in the survey are selected through a multistage sampling process. This method ensures that the sample accurately represents the population at national, urban-rural, and regional levels (such as counties or states).

The DHS surveys gather between 5,000 and 32,000 participants, who provide data through a standardized questionnaire covering various health indicators. These indicators include maternal demographics, household and community characteristics, child factors, and malnutrition. Detailed sampling techniques are available on the DHS website (https://www.dhsprogram.com/Methodology/Survey-Types/DHS-Methodology.cfm) (44) and literature (45).

The study includes 10 countries that have recorded data on female genital mutilation in their most recent DHS. The study population included 189155 women of reproductive age. This was reduced to 58,437 women who have reported whether their girl child has either undergone FGM or not. Due to the measures used to calculate the SWPER women empowerement variable, the data was further reduced to include 49,501 married or women cohabitating with their partners and having atleast one daughter age between 0 to 14 years, who has either undergone FGM or not (32). The data for this study is freely available at https://dhsprogram.com/Data/. Table 1 shows the countries, the survey years and weighted samples.

**Table 1:** Countries, survey year and sample included in the study.

| Country | Survey year | Study Sample |  |  |  |
| --- | --- | --- | --- | --- | --- |
|  |  | Non-weighted | Percent (%) | Weighted | Percent (%) |
| Burkina Faso | 2021 | 8,796 | 17.77 | 8,758 | 17.86 |
| Chad | 2014 - 15 | 4,818 | 9.73 | 4,727 | 9.64 |
| Gambia | 2019-20 | 2,541 | 5.13 | 2,300 | 4.69 |
| Guinea | 2018 | 5,149 | 10.4 | 5,068 | 10.33 |
| Kenya | 2022 | 6,584 | 13.3 | 6,217 | 12.67 |
| Mali | 2021 | 2,633 | 5.32 | 2,896 | 5.90 |
| Mauritania | 2019 - 20 | 2,423 | 4.89 | 2,462 | 5.02 |
| Nigeria | 2018 | 7,670 | 15.49 | 7,813 | 15.93 |
| Sierra Leone | 2019 | 6,249 | 12.62 | 6,165 | 12.57 |
| Tanzania | 2022 | 2,638 | 5.33 | 2,645 | 5.39 |
| **Total** |  | **49,501** |  | **49,050** |  |

### Study Variables

The outcome variable of this study is FGM of a daughter of a mother included in the study. this variable is available in the women questionnaire of the DHS dataset for some selected countries. The variable was coded as 0 “no FGM” and 1 “FGM”.

The main predictor variable in this study is women empowerment which was measured using the SWPER Global (46). The SWPER Global is a carefully designed women empowerement measure using the DHS for 62 lower-and middle-income countries (32, 46). The SWPER Global index consist of three key domains; i.Attitude towards violence domain, ii. Social Independence domain, and iii. Decision making domain based on 14 questions from the DHS women questionnaire.

The attitude towards violence domain assess a woman’s opinion regarding husband beating wife is justified in some specific situations. The social independence domain assesses woman’s access to information, attaining diserable educational level, age at marriage and first child, and also the differences in age and education to the cohabiting partner. The decision making domain measures who makes decisions in the household and the woman’s works. Women were categorized under low empowerment, medium empowerment, and high empowerment based on the cutoff points in the methodology of SWPER Global. The design of this index is comprehensively presented by Ewerling and colleagues (32, 46). The STATA do.file for calculating the SWPER index is available at https://goo.gl/isGonn (32).

A fourth “combined domain” was constructed to measure the cumulative empowerment of women using all three domains provided by Ewerling et al., (2020) and its association with FGM. Principal Component Analysis (PCA) was used to calculate the composite scores of all three domains considering the levels of empowerment scores in each domains recorded among women in this study. The STATA code used to arrive at the combined domain is presented in supplementary file 1. It should be used in addition to the do.file provided here https://goo.gl/isGonn (32). The combined domain is also measured using low, medium and high empowerement levels.

Other variables such as maternal age, maternal education, religion, wealth index, place of residence, FGM status of mother, partner’s age, partner’s educational level, partners occupation, and sex of household head were included in the study as covariates.

### Data Analysis

The study involved four levels of analysis. The first level was to calculate the levels of women empowerment among women in all four domains. This was done using descriptive statistics. The second level of the analysis involved Pearson’s chi-square test to assess the association between women empowerment and FGM at a significance level of p ≤ 0.05. Also, the association between each country and the level of empowerments and prevalence of FGM.

The variables significantly associated with FGM at p-value ≤ 0.05 were included in a regression model to ascertain the predictors of FGM. Bivariate logistric regression model (crude odds ratio) was used to examine the individual predictors of FGM, starting with SWPER women empowerment domains, and then the other covariates. All variables, SWPER index and covariates, significant at p-value ≤ 0.05 at a 95% confidence interval (CI), the bivariate regression model were included in a multivariate logistic regression, adjusting for confounders.

The data was analysed using STATA version 18 macOS. The analysis was performed considering the complex survey design implemented by DHS program. The STATA command “svy” was used in addition with the typical commands to account for the weighted data. The dumbell plots were designed using ggplot in R-studio, the cluster graphs designed using Microsoft excel version 16.87 and ArcMap version 10.2 used to design the symbology map.

Variance inflation factor (VIF) was used to test the multicollinearity of the study variables. There was no evidence of multicollinearity (mean vif = 1.65, min = 1.02, and max = 2.77).

## Results

A high percentage of individuals with high empowerment (47.96%) exhibit a positive attitude towards violence. Conversely, in the social independence domain, many of the mothers had low empowerment (48.32%). The distribution of these attributes is fairly balanced across all empowerment levels in the decision taking domain, with a slight predominance in the high empowerment group (33.58%). There was an almost equal distribution of levels of empowerment in the combined domain, but with a little surge in the low empowerment (35.63%). This is illustrated in Figure 1.

**Figure 1:**
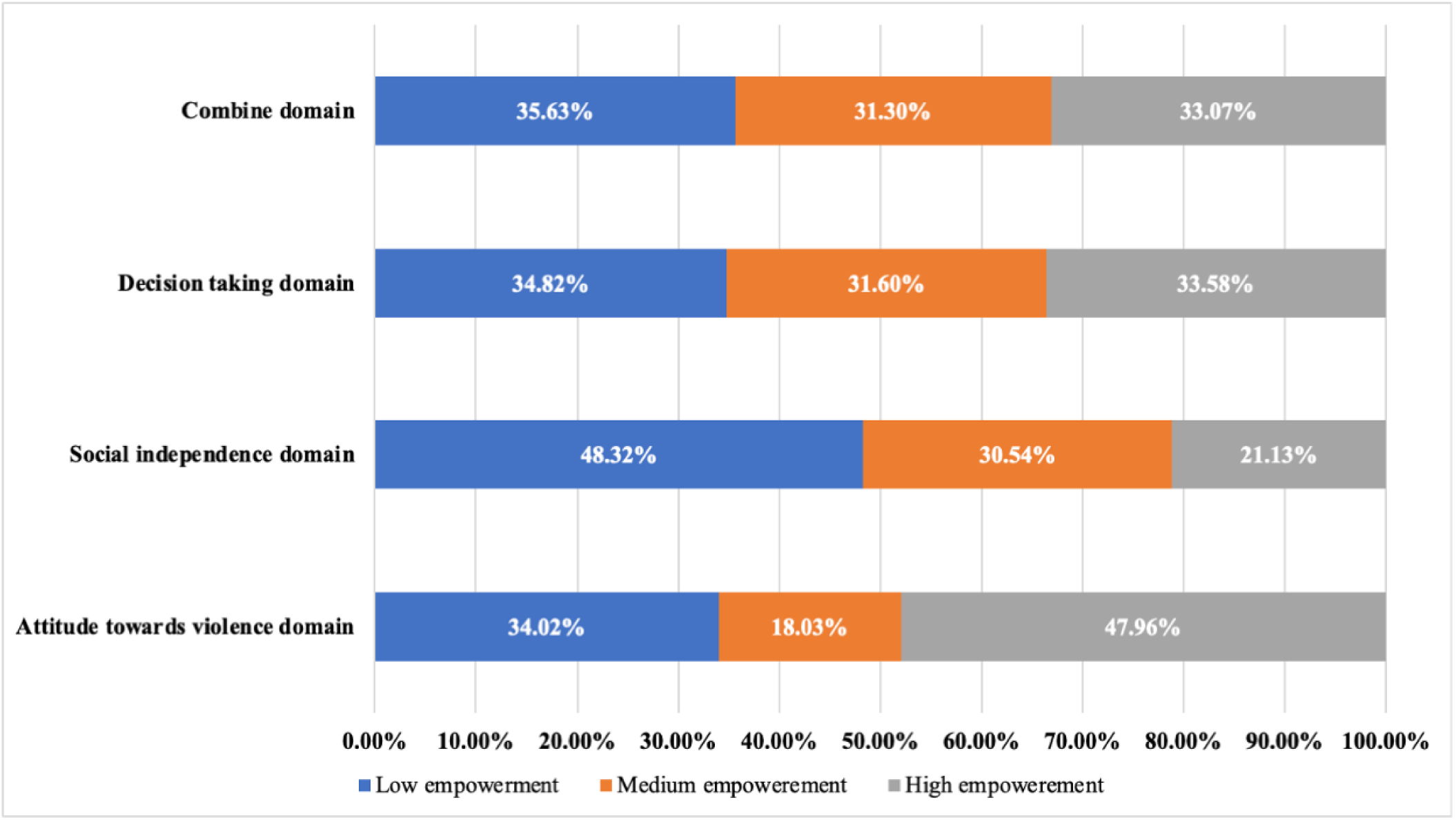
Levels of women empowerment in SWPER Domain

There is a significantly lower percentage of FGM among girls with mothers having high empowerement under the attitude towards violence domain (15.6%) compared to those with lower empowerement (22.59%) (χ2 = 330.4, p < 0.0001). Under the social independence, higher empowerment is strongly associated with a lower percentage of FGM cases (11.07%), compared to 23.30% FGM rate with mothers with lower empowerement (χ2 = 858.6, p < 0.0001). Also, under the decision taking domain, mothers with higher empowerement is associated with a lower (13.49%) FGM rate (χ2 = 512, p < 0.0001). In the combined domain, 23.34% of the mothers with high empowerment had girls with FGM compared to 46.88% of mothers with low empowerment (Table 2).

**Table 2:** Women Empowerement and its association with FGM.

| SWERP Domain | Empowerment level | FGM | | $\chi^2$ | p- value |
| --- | --- | --- | --- | --- | --- |
|  |  | No (%) | Yes (%) |  |  |
| **Attitude towards violence domain** | Lower empowerment | 12915 (77.41) | 3769 (22.59) | 330.4 | p < 0.0001 |
|  | Medium empowerment | 7319 (82.77) | 1524 (17.23) |  |  |
|  | High empowerment | 19853 (84.40) | 3670 (15.60) |  |  |
| **Social independence domain** | Lower empowerment | 18179 (76.70) | 5524 (23.30) | 858.6 | p < 0.0001 |
|  | Medium empowerment | 12690 (84.70) | 2291 (15.30) |  |  |
|  | High empowerment | 9219 (88.93) | 1148 (11.07) |  |  |
| **Decision taking domain** | Lower empowerment | 13149 (77.00) | 3928 (23.00) | 512.895 | p < 0.0001 |
|  | Medium empowerment | 12690 (81.86) | 2813 (18.14) |  |  |
|  | High empowerment | 14249 (86.51) | 2222 (13.49) |  |  |
| **Combined domains** | Lower empowerment | 13501 (33.19) | 4135 (46.88) | 696.6 | p < 0.0001 |
|  | Medium empowerment | 12869 (31.63) | 2627 (29.78) |  |  |
|  | High empowerment | 14310 (35.18) | 2059 (23.34) |  |  |

Countries like Tanzania (52.31%), Nigeria (69.22%), Mauritania (68.72%) and Kenya (59.89%) had high proportion of maternal empowerment against attitude towards violence, unlike Mali (58.18%), Guinea (56.01%) and Chad (51.52%) which showed very low maternal empowerment against attitude towards violence. Regarding social independence, most of the women in all the countries exhibited low empowerment levels. Most of the mothers in Kenya (65.28%), Mauritania (56.54%) and Tanzania (57.32%) had high empowerment level under the decision making domain, and mothers in Mali (63.96%) had very low empowerment. All the three domains had significantly different levels of empowerment between the countries (Attitude towards violence [χ2= 600.21, p < 0.0001], social independence [χ2 = 4301.3, p < 0.0001], and decision making [χ2 = 8201.1, p < 0.0001]). This is presented in figure 2.

**Figure 2:**
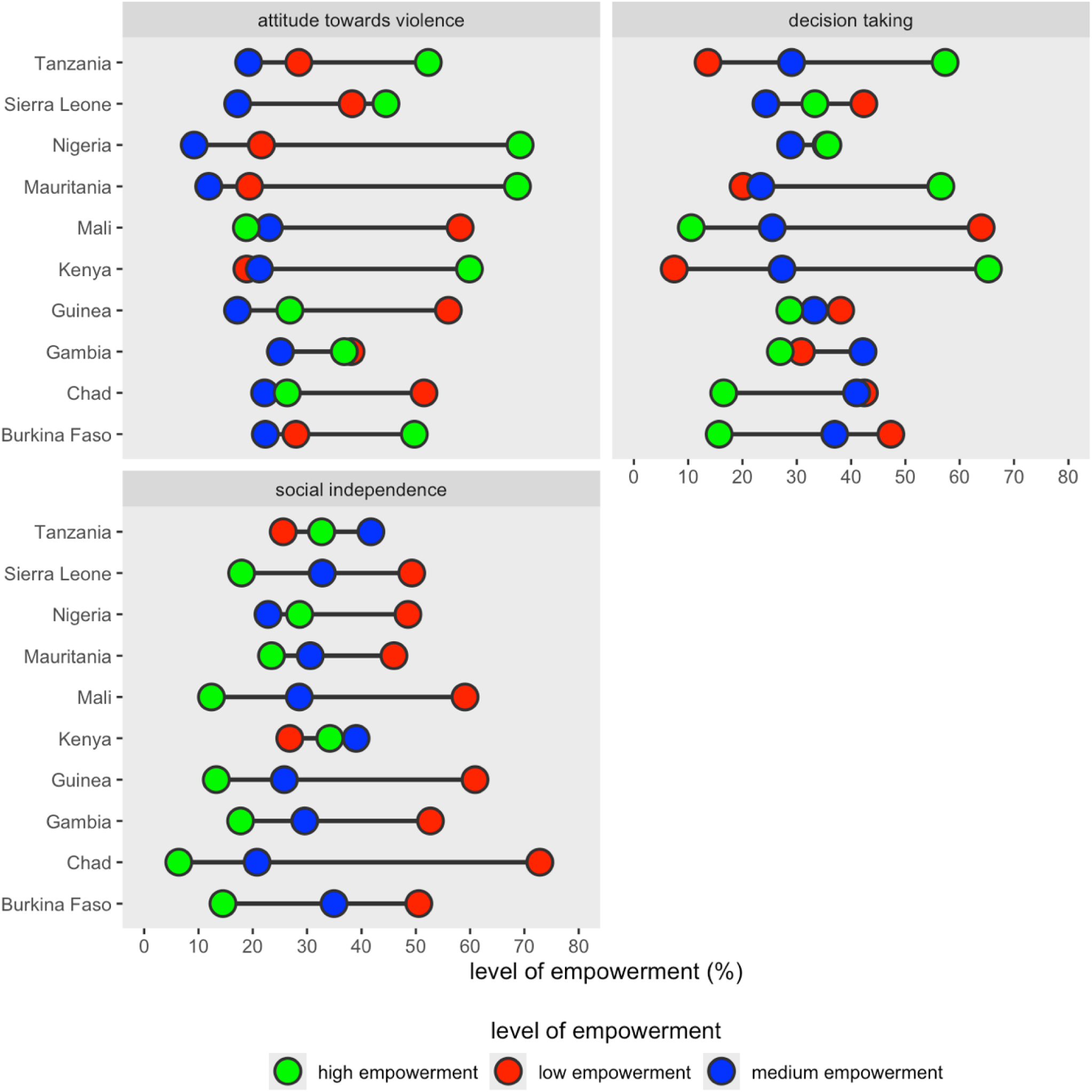
Women empowerment by countries

When all the levels of empowerment were combined, many of the women in Kenya (42.98%), Chad (39.89%)| and Tanzania (34.95%) had high level of empowerment, unlike most of the countries. Mali (52.41%), Guinea (48.86%) and Gambia (42.23%) showed majority of their women having low empowerment levels. There was a significant association between the countries and the levels of empowerment (χ2 = 1000.6, p < 0.0001). This is shown in figure 3.

**Figure 3:**
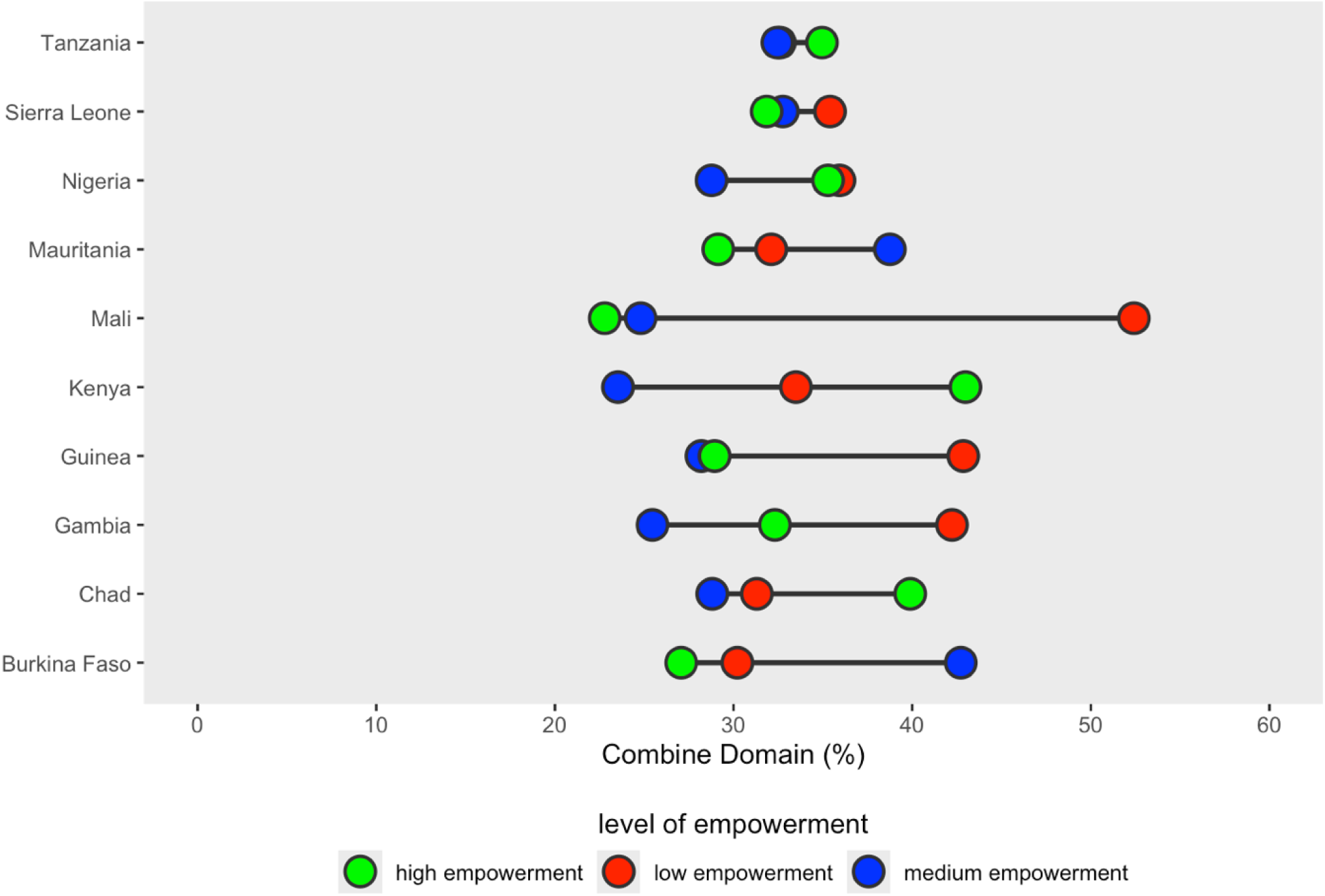
Combine Domain for SWPER Global index

Figure 4 shows the spatial distribution of high empowered mothers under the total empowerment domain and the prevalence of FGM in countries within SSA. Tanzania and Kenya had high empowerment levels of 34.95% and 42.98%, respectively, with very low FGM rates (Kenya = 2.35% and Tanzania = 0.34%) compared to countries like Mali and Gambia with very low empowerement levels (22.79% and 32.31%, respectively) and high prevalence of FGM (Mali = 61.3%, Gambia = 40.42%). Also, Nigeria had relatively high maternal empowerement (35.2%) and equally high FGM rates (26.48%). There was a significant association between countries and the prevalence of FGM (χ2 = 9100, p < 0.0001) and total women empowerement (χ2 = 1600, p < 0.0001).

**Figure 4:**
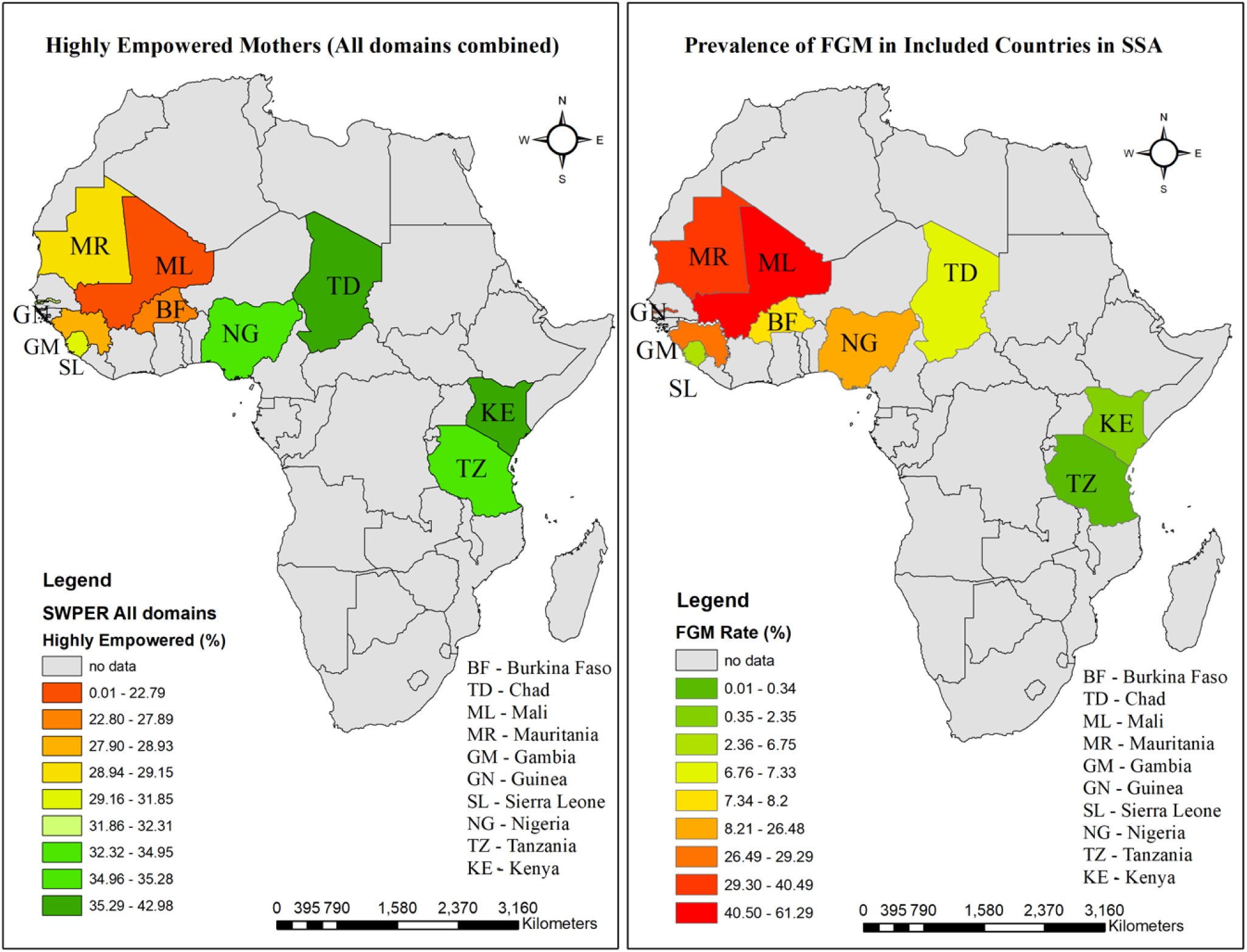
High empowerment level in the combine domain and FGM by countries

Many of the mothers included in the study were age between 25 – 29 years (21.81%) and 30 – 34 years (21.07%). Mothers belonging to the Islamic religion were 47.32%. Also, many of the mothers were in the poorest wealth index (21.11%) and majority living in rural areas (66.82%). Majority of the mothers included in the study were circumcised (61.01%). Almost half (49.68%) of the mothers had partners age 40 – 59 years, with 52.33% of them not having formal education. Majority of them had male family heads (86.21%). The prevalence of FGM among girls increases with age of mothers peaking at 27.60% in the 45 – 49 age group (χ2 = 388.35, p < 0.0001). There was also a significantly high prevalence of FGM among girls with Muslim mothers (29.58%) compared to Christians (4.88%) (χ2 = 4,483.22, p < 0.0001). Maternal education was associated with high FGM prevalence among girls, with 23.86% (χ2 = 1313.44, p < 0.0001) among those with no education. Maternal circumcision was highly significantly associated with FGM of girl child (χ2 = 3802.3, p < 0.0001), with circumcised mothers having more female children who were also circumcised (26.83%), compared to 4.88% of mothers not circumcised. Older and less educated partners were also found to be associated with higher FGM rates (p < 0.0001). These are presented in table 3.

**Table 3:** Association between other confounding variables and FGM.

| Socio-demographic Characteristics | Frequency<br>(n = 49,501) | Percent<br>(%) | Female Genital Mutilation | | $\chi^2$ | p- value |
| --- | --- | --- | --- | --- | --- | --- |
|  |  |  | No (%) | Yes (%) |  |  |
| **Age** |  |  |  |  | 388.35 | < 0.0001 |
| 15-19 | 1466 | 2.96 | 1217 (83.77) | 236 (16.23) |  |  |
| 20-24 | 6210 | 12.55 | 5172 (84.05) | 982 (15.95) |  |  |
| 25-29 | 10797 | 21.81 | 9029 (84.39) | 1670 (15.61) |  |  |
| 30-34 | 10430 | 21.07 | 8573 (82.95) | 1762 (17.05) |  |  |
| 35-39 | 9780 | 19.76 | 7951 (82.04) | 1741 (17.96) |  |  |
| 40-44 | 6528 | 13.19 | 5069 (78.36) | 1400 (21.64) |  |  |
| 45-49 | 4290 | 8.67 | 3078 (72.40) | 1173 (27.60) |  |  |
| **Religion** |  |  |  |  | 4,483.22 | < 0.0001 |
| Christianity | 19945 | 40.29 | 18799 (95.12) | 965 (4.88) |  |  |
| Islam | 23423 | 47.32 | 16345 (70.42) | 6865 (29.58) |  |  |
| No region | 378 | 0.76 | 300 (80.11) | 74 (19.89) |  |  |
| other | 581 | 1.17 | 531 (92.19) | 45 (7.81) |  |  |
| Missing | 5173 | 10.45 |  |  |  |  |
| **Educational status** |  |  |  |  | 1,313.44 | < 0.0001 |
| no education | 27274 | 55.1 | 20577 (76.14) | 6449 (23.86) |  |  |
| primary | 10591 | 21.4 | 9258 (88.22) | 1236 (11.78) |  |  |
| secondary | 9205 | 18.6 | 8003 (87.73) | 1119 (12.27) |  |  |
| higher | 2431 | 4.91 | 2249 (93.40) | 159 (6.60) |  |  |
| **Wealth index** |  |  |  |  | 371.32 | < 0.0001 |
| poorest | 10451 | 21.11 | 8024 (77.49) | 2331 (22.51) |  |  |
| poorer | 10163 | 20.53 | 8037 (79.80) | 2034 (20.20) |  |  |
| middle | 10116 | 20.44 | 8138 (81.19) | 1885 (18.81) |  |  |
| richer | 9654 | 19.5 | 7991 (83.53) | 1575 (16.47) |  |  |
| richest | 9117 | 18.42 | 7897 (87.41) | 1137 (12.59) |  |  |
| **Place of residence** |  |  |  |  | 99.85 | < 0.0001 |
| urban | 16422 | 33.18 | 13700 (84.19) | 2573 (15.81) |  |  |
| rural | 33079 | 66.82 | 26388 (80.50) | 6390 (19.50) |  |  |
| **FGM status** |  |  |  |  | 3,802.30 | < 0.0001 |
| Not circumcised | 19300 | 38.99 | 18192 (95.12) | 932 (4.88) |  |  |
| circumcised | 30200 | 61.01 | 21895 (73.17) | 8030 (26.83) |  |  |
| **Partner's age** |  |  |  |  | 755.04 | < 0.0001 |
| less than 20 | 175 | 0.35 | 150 (86.23) | 24 (13.77) |  |  |
| 21 - 39 | 20499 | 41.41 | 17638 (86.84) | 2674 (13.16) |  |  |
| 40 - 59 | 24593 | 49.68 | 19273 (79.09) | 5096 (20.91) |  |  |
| 60 and above | 4170 | 8.42 | 2963 (71.70) | 1169 (28.30) |  |  |
| Missing | 64 | 0.13 |  |  |  |  |
| **Partner's educationa level** |  |  |  |  | 1470.36 | < 0.0001 |
| education | 25904 | 52.33 | 19361 (75.43) | 6307 (24.57) |  |  |
| primary | 8931 | 18.04 | 7984 (90.21) | 866 (9.79) |  |  |
| secondary | 10349 | 20.91 | 8912 (86.90) | 1343 (13.10) |  |  |
| higher | 4252 | 8.59 | 3767 (89.40) | 447 (10.60) |  |  |
| Missing | 64 | 0.13 |  |  |  |  |
| **Partner's occupation** |  |  |  |  | 725.39 | < 0.0001 |
| not working | 3452 | 6.97 | 2724 (79.63) | 697 (20.37) |  |  |
| managerial | 4778 | 9.65 | 4162 (87.89) | 573 (12.11) |  |  |
| clerical | 533 | 1.08 | 404 (76.44) | 124 (23.56) |  |  |
| sales | 7367 | 14.88 | 5515 (75.54) | 1786 (24.46) |  |  |
| agricultural | 19451 | 39.29 | 15811 (82.04) | 3462 (17.96) |  |  |
| other | 1496 | 3.02 | 1127 (76.00) | 356 (24.00) |  |  |
| services | 3215 | 6.5 | 2341 (73.47) | 845 (26.53) |  |  |
| manual | 9144 | 18.47 | 7941 (87.65) | 1119 (12.35) |  |  |
| Missing | 64 | 0.13 |  |  |  |  |
| **Sex of family head** |  |  |  |  | 17.71 | < 0.01 |
| male | 42675 | 86.21 | 34436 (81.44) | 7850 (18.56) |  |  |
| female | 6826 | 13.79 | 5651 (83.56) | 1112 (16.44) |  |  |

### Multivariate Logistic Regression Models

Table 4 shows the results of the logistic regression models. The level of empowerement in all SWPER domains were found to be significant predictors of FGM, with medium and high empowered mothers having significantly lower odds of child with FGM in the unadjusted model (p < 0.0001). However, after adjusting for confounders, mothers with high empowerment under the attitude towards violence were 13% more likely to have a daughter with FGM (AOR: 1.130, 95%CI: 1.038 – 1.123)). Under the social independence domain, high empowered mothers were 27.9% more less likely to have child with FGM (AOR: 0.721; 95%CI: 0.648 – 0.802). Similarly, women who were medium empowered regarding decision taking were 10.2% less likely to have child with FGM compared to those having low empowerement. Mothers with high empowerment in all the domains were over 50% less likely to have a daughter with FGM (AOR: 0.583, 95%CI: 0.523 – 0.649).

**Table 4:**
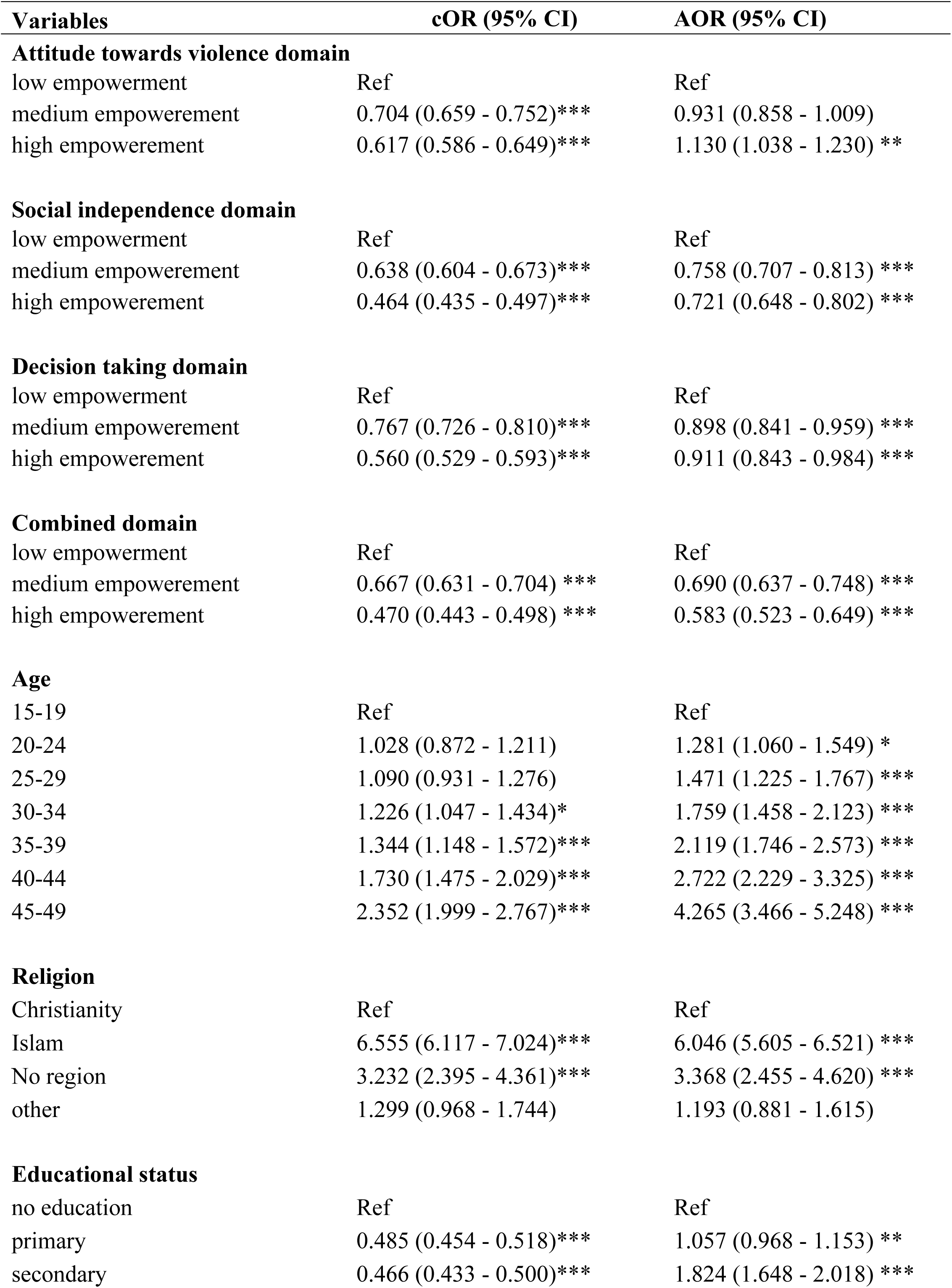
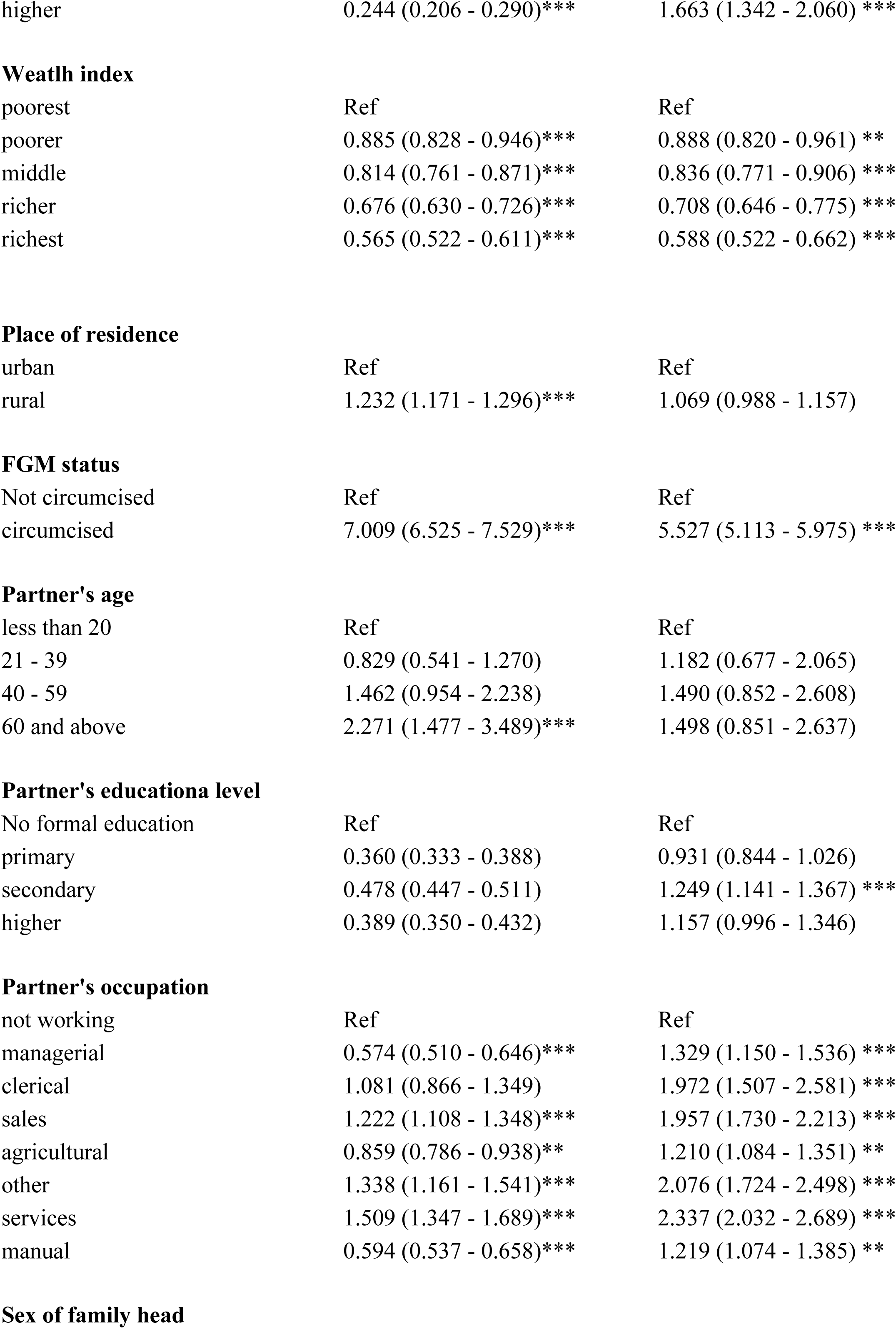
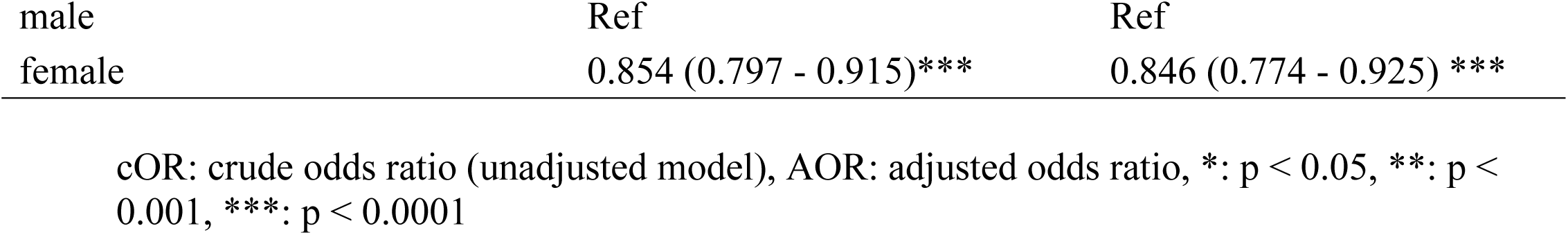
Multivariate logistic regression model on SWPER model and FGM.

| Variables | cOR (95% CI) | AOR (95% CI) |
| --- | --- | --- |
| **Attitude towards violence domain** |  |  |
| low empowerment | Ref | Ref |
| medium empowerment | 0.704 (0.659 - 0.752)*** | 0.931 (0.858 - 1.009) |
| high empowerment | 0.617 (0.586 - 0.649)*** | 1.130 (1.038 - 1.230) ** |
| **Social independence domain** |  |  |
| low empowerment | Ref | Ref |
| medium empowerment | 0.638 (0.604 - 0.673)*** | 0.758 (0.707 - 0.813) *** |
| high empowerment | 0.464 (0.435 - 0.497)*** | 0.721 (0.648 - 0.802) *** |
| **Decision taking domain** |  |  |
| low empowerment | Ref | Ref |
| medium empowerment | 0.767 (0.726 - 0.810)*** | 0.898 (0.841 - 0.959) *** |
| high empowerment | 0.560 (0.529 - 0.593)*** | 0.911 (0.843 - 0.984) *** |
| **Combined domain** |  |  |
| low empowerment | Ref | Ref |
| medium empowerment | 0.667 (0.631 - 0.704) *** | 0.690 (0.637 - 0.748) *** |
| high empowerment | 0.470 (0.443 - 0.498) *** | 0.583 (0.523 - 0.649) *** |
| **Age** |  |  |
| 15-19 | Ref | Ref |
| 20-24 | 1.028 (0.872 - 1.211) | 1.281 (1.060 - 1.549) * |
| 25-29 | 1.090 (0.931 - 1.276) | 1.471 (1.225 - 1.767) *** |
| 30-34 | 1.226 (1.047 - 1.434)* | 1.759 (1.458 - 2.123) *** |
| 35-39 | 1.344 (1.148 - 1.572)*** | 2.119 (1.746 - 2.573) *** |
| 40-44 | 1.730 (1.475 - 2.029)*** | 2.722 (2.229 - 3.325) *** |
| 45-49 | 2.352 (1.999 - 2.767)*** | 4.265 (3.466 - 5.248) *** |
| **Religion** |  |  |
| Christianity | Ref | Ref |
| Islam | 6.555 (6.117 - 7.024)*** | 6.046 (5.605 - 6.521) *** |
| No region | 3.232 (2.395 - 4.361)*** | 3.368 (2.455 - 4.620) *** |
| other | 1.299 (0.968 - 1.744) | 1.193 (0.881 - 1.615) |
| **Educational status** |  |  |
| no education | Ref | Ref |
| primary | 0.485 (0.454 - 0.518)*** | 1.057 (0.968 - 1.153) ** |
| secondary | 0.466 (0.433 - 0.500)*** | 1.824 (1.648 - 2.018) *** |
| higher | 0.244 (0.206 - 0.290)*** | 1.663 (1.342 - 2.060) *** |
| Wealth index |  |  |
| poorest | Ref | Ref |
| poorer | 0.885 (0.828 - 0.946)*** | 0.888 (0.820 - 0.961) ** |
| middle | 0.814 (0.761 - 0.871)*** | 0.836 (0.771 - 0.906) *** |
| richer | 0.676 (0.630 - 0.726)*** | 0.708 (0.646 - 0.775) *** |
| richest | 0.565 (0.522 - 0.611)*** | 0.588 (0.522 - 0.662) *** |
| Place of residence |  |  |
| urban | Ref | Ref |
| rural | 1.232 (1.171 - 1.296)*** | 1.069 (0.988 - 1.157) |
| FGM status |  |  |
| Not circumcised | Ref | Ref |
| circumcised | 7.009 (6.525 - 7.529)*** | 5.527 (5.113 - 5.975) *** |
| Partner's age |  |  |
| less than 20 | Ref | Ref |
| 21 - 39 | 0.829 (0.541 - 1.270) | 1.182 (0.677 - 2.065) |
| 40 - 59 | 1.462 (0.954 - 2.238) | 1.490 (0.852 - 2.608) |
| 60 and above | 2.271 (1.477 - 3.489)*** | 1.498 (0.851 - 2.637) |
| Partner's educationa level |  |  |
| No formal education | Ref | Ref |
| primary | 0.360 (0.333 - 0.388) | 0.931 (0.844 - 1.026) |
| secondary | 0.478 (0.447 - 0.511) | 1.249 (1.141 - 1.367) *** |
| higher | 0.389 (0.350 - 0.432) | 1.157 (0.996 - 1.346) |
| Partner's occupation |  |  |
| not working | Ref | Ref |
| managerial | 0.574 (0.510 - 0.646)*** | 1.329 (1.150 - 1.536) *** |
| clerical | 1.081 (0.866 - 1.349) | 1.972 (1.507 - 2.581) *** |
| sales | 1.222 (1.108 - 1.348)*** | 1.957 (1.730 - 2.213) *** |
| agricultural | 0.859 (0.786 - 0.938)** | 1.210 (1.084 - 1.351) ** |
| other | 1.338 (1.161 - 1.541)*** | 2.076 (1.724 - 2.498) *** |
| services | 1.509 (1.347 - 1.689)*** | 2.337 (2.032 - 2.689) *** |
| manual | 0.594 (0.537 - 0.658)*** | 1.219 (1.074 - 1.385) ** |
| Sex of family head |  |  |
| male | Ref | Ref |
| female | 0.854 (0.797 - 0.915)*** | 0.846 (0.774 - 0.925) *** |

cOR: crude odds ratio (unadjusted model), AOR: adjusted odds ratio, \*: $p < 0.05$ , \*\*: $p < 0.001$ , \*\*\*: $p < 0.0001$

It was also observed that increasing age was signficantly associated with higher odds of FGM, with mothers in 40 – 44 age group having more than twice higher odds (AOR: 2.722, 95%CI: 2.229 – 3.325) compared to those in 15 – 19 age group. Mothers in the Islamic religion had 6 times higher odds of having child with FGM (AOR: 6.046; 95%CI: 5.605 – 6.521) compared to Christian mothers. Increasing wealth index was also found to be a protective factor of FGM, with mothers in the richest wealth index having 42.1% lower odds (AOR: 0.588; 95%CI: 0.522 – 0.662). Mothers who were also circumcised were 5 times more likely to have their female child circumcised (AOR: 5.527; 95%CI: 5.113 – 5.975). Female household head was found to be a protective factors of FGM (AOR: 0.846; 95%CI: 0.774 – 0.925).

## Discussion

This study aimed to investigate the impact of women’s empowerment, using the SWPER model, on FGM in Sub-Saharan Africa (SSA). The study included 10 SSA countries that reported FGM in their national demographic health surveys. Nearly half of highly empowered individuals (47.96%) have a positive attitude towards violence. In the social independence domain, many mothers show low empowerment (48.32%). Empowerment levels in decision-making and the combined domain were fairly balanced, with many in high empowerment group (33.58%) for the decision making domain and many in the lower empowerment group (35.63%) in the combined domain. The prevalence of FGM was disproportionately distributed across the countries included in the study. The levels of empowerment in all of the domains and the prevalence of FGM were significantly associated with the countries. Countries such as Mali (61.3%), and Gambia (40.42%), located in Western Africa, reported higher rates of FGM, while Kenya (2.35%) and Tanzania (0.34%), located in East Africa, reported lower FGM rates.

The overall rate of FGM in SSA has seen a decline over the years (13, 47). Several countries such as Tanzania and Kenya, with low rates of FGM have made significant strides in women’s empowerment and education, leading to a cultural shift away from FGM (42, 48, 49). Community-based programs and legal frameworks have also played a role in reducing FGM rates (50). In contrast, some countries in the West of SSA still shows a strong cultural and traditional support for FGM, making it harder to eradicate despite efforts to empower women (13, 19). Countries such as Nigeria has diverse cultural landscape includes regions with strong traditional support for FGM. Empowerement efforts may not be uniformly effective across all regions, leading to higher rates in certain areas (51). Targetted interventions are therefore required in the effort to eradicate the practice in such diverse regions

The findings showed that nearly half of highly empowered individuals have a positive attitude towards violence. This reflect the complex socio-cultural dynamics in SSA. In many communities, traditional norms and practices, including certain forms of violence, are deeply ingrained. The high percentage of mothers with low social independence empowerment (48.32%) highlights the challenges women face in SSA. Social norms often restrict women’s autonomy and mobility, limiting their ability to make independent decisions. This lack of social independence can perpetuate practices like FGM, as women may feel pressured to conform to community expectations. The fairly balanced empowerment levels in decision-making, with a slight majority in the high empowerment group (33.58%), suggest that some progress is being made in SSA. However, the slight predominance indicates that many women still struggle to assert their decision-making power fully. Cultural expectations and patriarchal structures often limit women’s roles in household and community decisions. Practices such as FGM are deeply rooted in cultural and religious beliefs systems in SSA (19, 52).

In this study, highly empowered mothers were less likely to have a child with FGM. This suggests that empowerment in rejecting violence correlates with lower FGM rates, possibly due to increased awareness and rejection of harmful practices. This finding reflects in an earlier study in SSA assessing how women empowerement and attitude towards violence can influence their intention towards FGM (41). High empowerment in social independence led to a 41.2% reduction in FGM odds. Empowered women may have better access to resources and support networks, enabling them to resist FGM. Medium empowerment in decision-making resulted in reduction in FGM odds. This suggest that women with decision-making power can better protect their children from FGM. Decision making forms a critical component of women ability to accept or decline certain practices or healthcare interventions. Other studies emphasized the important role a woman’s ability to take independent decision positively influence certain health outcomes.

Older mothers (45–49) had more than four times higher odds of having a child with FGM. This could be due to generational differences in attitudes towards FGM, with older mothers more likely to adhere to traditional practices. This finding corresponds with earlier studies in the SSA region where the practice of FGM was still common among older mothers (19, 40, 53). Younger generations and more empowered women may be driving cultural shifts away from traditional practices like FGM.

Mothers who were circumcised were five times more likely to have their female child circumcised. This indicates a cycle of perpetuation, where circumcised mothers continue the practice with their daughters. Both similar (54) and contrasting finding (39) has been reported in an earlier studies. It is important to implement educational initiatives that focus on women’s rights, health, and the dangers of FGM. Empower women through knowledge and skills training. This can help break the circle of perpetuation.

Mothers in the Islamic religion had almost six times higher odds of having a child with FGM compared to Christian mothers. This may reflect cultural or religious norms that support FGM in certain communities. This study finding corresponds with other studies in the SSA sub-region (39, 43). Other researcher argue that, although the practice of FGM may be common among Muslims, it is more of a cultural practice since it is not mandated in the religion (55). Nonetheless, to eradicate this practice, it is essential to engage both religious and traditional leaders sensitization programs and to speak out against the practice.

Increasing wealth was protective against FGM, with the richest mothers having 46.9% lower odds. Wealthier families may have better access to education and healthcare, reducing the prevalence of FGM. This finding is supported in earlier studies (39–41). Women with greater economic independence are more likely to challenge traditional norms and practices. Also, empowered and wealthier women may have better access to resources and support systems that help them resist societal pressures to perform FGM.

Female household heads were found to be a protective factor against FGM. Women leading households may have more autonomy and decision-making power to prevent FGM. It will be helpful to encourage women to take on leadership roles within their communities to influence change and advocate against FGM.

## Conclusion

This study highlights the intricate relationship between women’s empowerment and the prevalence of FGM in SSA. The findings suggest that higher levels of empowerment, particularly in rejecting violence and decision-making, are associated with lower rates of FGM. There was also a strong association between levels of empowerment in the various domains and the prevalence of FGM across the countries. Despite progress in some regions, traditional norms and cultural support for FGM persist, especially in Western Africa, posing challenges to eradication efforts. The study presents the importance of targeted empowerment initiatives that address socio-cultural dynamics and promote women’s autonomy to continue reducing FGM prevalence across SSA.

Education is seen as a powerful tool to eradicating FGM. Increasing access to education, women and girls can gain the knowledge and skills required to overcome the challenges of the practice. Additionally, enactment, enforcement and strengthening legal frameworks that prohibit FGM can provide a strong deterrent against the practice. The legal frameworks should be supported by community awareness campaigns, spearheaded by traditional and religious leaders, to ensure widespread understanding and compliance. Additionally, Community-based programs that involve local leaders and stakeholders can address the cultural and traditional norms that perpetuate FGM. Empowering women at the community level can lead to sustainable change. These efforts will promote the efforts of eradicating FGM in SSA, and achieving gender equality and empowerement of women and girls, as enshrined in the United Nations SDG 5.

## Declarations

### Ethical Consideration

This research used the DHS database, a global survey spanning five years. Permission to access and use the data was granted by ICF International following topic registration and submission on their website. Detailed methodology and ethical considerations are available on the DHS website. Since the data were secondary and publicly accessible, individual consent was not required by the ICF International Institutional Review Board. The study received all necessary approvals from the DHS Program, and data privacy was rigorously maintained during processing and analysis.

### Consent for Publication

Not applicable

### Availability of data and materials

The data for this study is freely available in the DHS website (https://dhsprogram.com/Data/).

### Authors’ Contribution

**Conceptualization** - MHK, JM; Methodology – MHK; Data Curation: MHK; Formal analysis – MHK; Writing of Original draft – JM; Writing review & editing – JM, Supervision & Validation – MHK.

## Acknowledgements

We would like to thank the Measure DHS Program for providing the DHS datasets.

